# The Measurement and Mitigation of Algorithmic Bias and Unfairness in Healthcare AI Models Developed for the CMS AI Health Outcomes Challenge

**DOI:** 10.1101/2022.09.29.22280537

**Authors:** Carol J. McCall, Dave DeCaprio, Joseph Gartner

## Abstract

Algorithms play an increasingly prevalent role in healthcare, and are used to target interventions, reward performance, and distribute resources, including funding. Yet it is widely recognized that many algorithms used today may inadvertently encode and perpetuate biases and contribute to health inequities. Artificial intelligence algorithms, in addition to being assessed for accuracy, must be evaluated with respect to whether they could impact disparities in health outcomes.

This paper presents details and results of ClosedLoop’s methods to measure and mitigate bias in machine learning models that were the winning submission in the CMS AI Health Outcomes Challenge. The submission applied a comprehensive framework for assessing algorithmic bias and fairness and the development and application of a metric appropriate for real-world healthcare settings capable of being used to assess and reduce the presence and impact of unfairness.

The submission demonstrated precision and transparency in the comprehensive measurement of algorithmic bias from multiple sources, including data representativeness, subgroup validity, label choice, and feature bias. For feature bias, the submission made a detailed examination of feature selection and diversity, including evaluating the appropriateness of including race in algorithm development. It also demonstrated how fairness criteria could be used to adjust care management enrollment thresholds to mitigate unfairness.

Computational methods and measures exist that allow healthcare organizations to measure and mitigate algorithmic bias and fairness in models used in practical healthcare settings. It is possible for healthcare organizations to adopt policies and practices that enable them to design, implement, and maintain algorithms that are highly accurate, unbiased, and fair.

**Author summary:** AI has come of age through the alchemy of cheap parallel (cloud) computing combined with the availability of big data and better algorithms. Problems that seemed unconquerable a few years ago are being solved, at times with startling gains. AI has finally arrived in health care, where the stakes are high, and the complexity and criticality of issues can far outweigh other applications. AI’s arrival is good; organizations are confronting forces strong enough that they may only yield once AI is brought to bear. AI has started to play a central role in targeting care interventions, rewarding physician performance, and distributing resources, including funding.

Here’s the problem: If health care’s algorithms are biased — something that researchers at the Center for Applied Artificial Intelligence at the University of Chicago’s Booth School of Business have concluded — then AI solutions designed to drive better outcomes can make things worse. The good news is that these experts also said that algorithmic bias, while pervasive, is not inevitable. The key is to define the processes and tools that can help measure and address it. The work presented in this paper represent an important contribution to these tools and a real-world demonstration of results.

## Introduction

Algorithms play an increasingly prevalent role in healthcare, and are used to target interventions, reward performance, and distribute resources, including funding.[1,2] But today’s mainstream algorithms have well-documented limitations, including issues related to racial and socioeconomic variables that lead to systemic inaccuracies, can exacerbate disparities, or be exploited for financial gain.[3-5] In 2019, researchers revealed that an algorithm widely used for population health management dramatically underestimated the health needs of the sickest Black patients.[5] More recently, researchers at Chicago Booth’s Center for Applied Artificial Intelligence examined a wide range of healthcare algorithms and concluded that bias in healthcare algorithms is pervasive.[6] U.S. leaders have urged the Centers for Medicare & Medicaid Services (CMS) to & #x201C;consider the risk for algorithmic bias and its potential impact on health disparities outcomes.”[7]

On March 27, 2019, CMS announced the Artificial Intelligence (AI) Health Outcomes Challenge.[8] The multi-stage, two year Challenge attracted over 300 entrants, including some of the nation’s leading healthcare and technology organizations. Its aim was to 1) showcase ways to better predict and improve healthcare outcomes, 2) create & #x2018;explainable AI solutions that clinicians trust’, and 3) identify ways to address algorithmic bias. Stage 1 models needed to predict unplanned hospital admissions, skilled nursing facility (SNF) admissions, and/or adverse events within a 30-day forecast window. The target event could be a specific event or a composite of events, but there could only be one forecast per beneficiary and date. Stage 2 models needed to predict mortality over a 12-month period. Models were judged in terms of accuracy and explainability. Accuracy was assessed by measuring Area Under the Curve (AUC), Sensitivity at Low Alert Rates, and Calibration indices. Explainability was judged on the extent to which explanatory displays ensured clinicians, and by extension patients, would sufficiently understand the predictive model forecasts to make reasonable use of those forecasts in care decisions. In Stage 2, participants were also required to explain how they addressed and mitigated implicit algorithmic bias in their submissions and report any quantitative results supporting their explanation. No other requirements regarding bias submissions were defined or framed for participants.

On April 30, 2021, CMS announced that ClosedLoop.ai had won the Challenge. This paper presents the details and results of that portion of ClosedLoop’s Stage 2 submission focused on methods used to measure and mitigate bias in machine learning models. Details regarding ClosedLoop’s submission for predictive models, including outcome rates and model-specific performance statistics, are included in Supplemental Material 1. Other submission details are also available on ClosedLoop’s website.[9]

## Results

### Reducing algorithmic bias

#### Representativeness of data for model development

All models were built using medical claims data provided by CMS. ClosedLoop evaluated groups that were identifiable within the data, shown in Table 1.

**Table 1.** Data representativeness.

| Population Group | Percent of CMS Data | Percent of U.S. Adults |
| --- | --- | --- |
| Race / Ethnicity |  |  |
| Asian | 2.4% | 6.7%<sup>a</sup> |
| Black | 10.4% | 13.4%<sup>a</sup> |
| Hispanic | 2.7% | 16.5%<sup>a</sup> |
| North American Native | 0.6% | 1.9%<sup>a</sup> |
| Other & Unknown | 4.0% | ---- |
| White | 79.8% | 77.5%<sup>a</sup> |
| Female Sex | 52.2% | 51.3%<sup>a</sup> |
| Disabled Individuals | 24.3% | 22.6%<sup>b</sup> |
| Dual Eligible | 17.5% | 4.7%<sup>c</sup> |
| Urban / Rural<sup>d</sup> |  |  |
| Urban Center | 47.5% | 54.6% |
| Rural | 2.2% | 1.5% |

<sup>a</sup>2019 Census data
<sup>b</sup>2010 Census Data
<sup>c</sup>2018 Census Data and CMS Facts
<sup>d</sup>2013 Rural-Urban Continuous (RUCC) Codes

Data were tested for representativeness of populations to which the models might possibly be applied. By definition, the CMS data was representative of traditional Medicare beneficiaries covered between 2013-2017. However, the data did not represent the distribution of these groups within U.S. adults generally.

#### Assessment of subgroup validity

To evaluate whether the models performed well for each subgroup, ClosedLoop used Matthew’s Correlation Coefficient (MCC).[10] MCC is the correlation of the observed values versus the predicted labels and a more valid metric when comparing groups with different prevalences. The threshold for MCC was set at 5% to reflect a typical proportion of beneficiaries typically enrolled in a high-risk program.

The 30-day Adverse Events Model and the 12-Month Morality model were well calibrated across subgroups, with a calibration index of less than 0.001 (Supplemental Material 2). Some variation in performance by subgroup was expected. AUC was within 3.2% of the population average for all subgroups and MCC @ 5% was within 2.1%.

#### Assessment of label choice bias

ClosedLoop’s used an approach like that used by Obermeyer et al. and compared the model’s calibration when using its original outcome versus a commonly used measure of chronic disease burden, the Charlson Comorbidity Index. The model was well calibrated for the original outcome (Fig. 1, top left), but showed greater racial differences when compared to the Charlson Comorbidity Index (Fig. 1, bottom left). This bias was the opposite of what is usually observed, with Blacks at the high-risk threshold having a lower average Charlson Comorbidity Index than Whites. Because one of the specific events in the 30-day Adverse Events composite risk prediction is an ED visit, it was hypothesized that the difference could be related to racial differences in emergency room use. To test this, ClosedLoop repeated the analysis (Fig. 1, top & bottom right) using an altered model that removed emergency room visits from the outcome, which reduced the racial differences in chronic disease burden.

**Fig 1.**
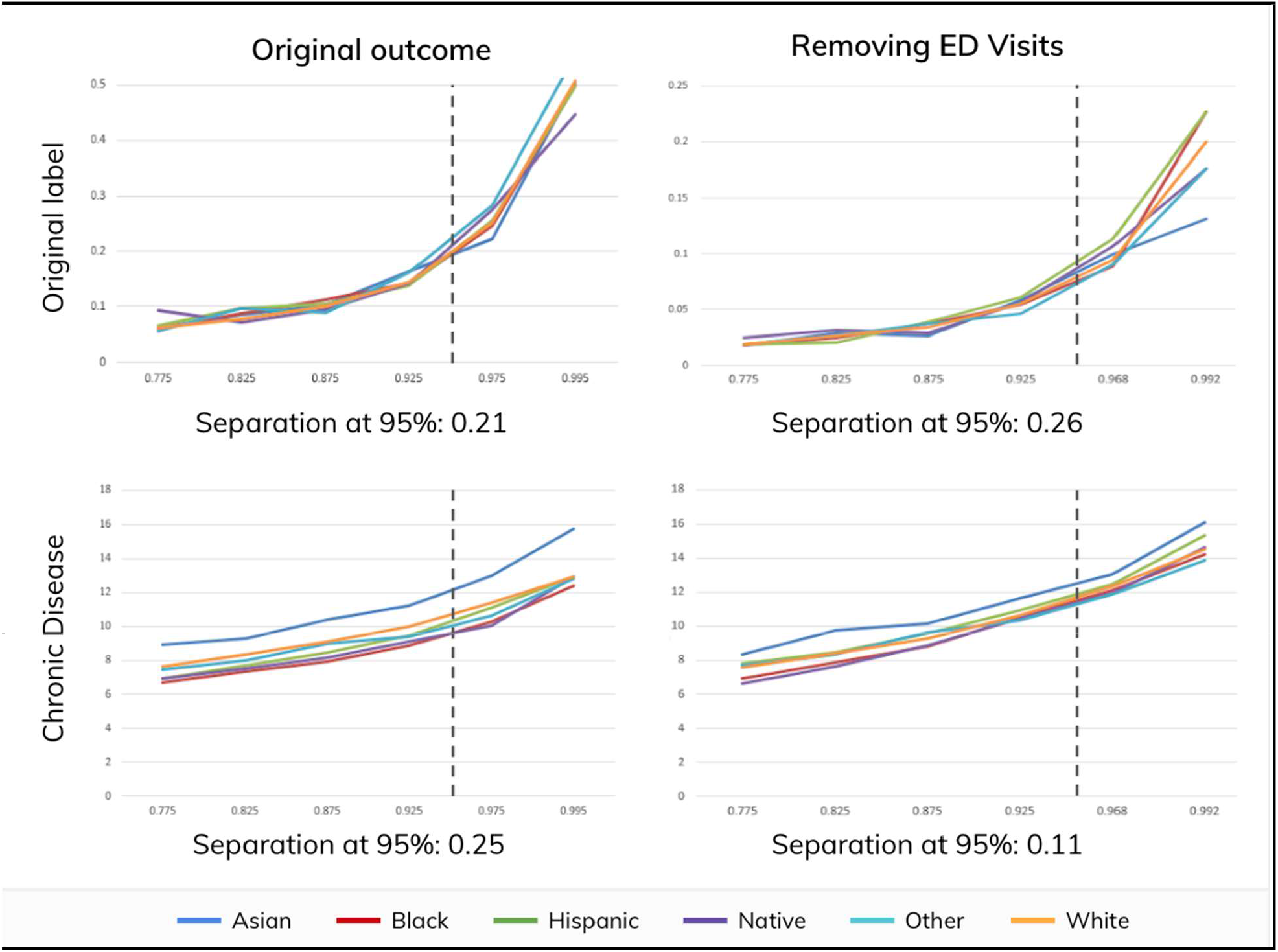
Assessment of label choice bias.

#### Assessment of feature bias

Models were developed from 1000s of candidate features. The final models included 753 features derived from claims and publicly available datasets (Supplemental Material 3). Several steps were taken to reduce or eliminate feature bias:

- Features were excluded that have been shown can perpetuate health disparities.[5] Cost features were not used anywhere in the models and the inclusion of race was guided by expert opinion.[11]
- Features were defined in ways that were robust to differences in data collection (e.g. counting distinct dates for diagnoses rather than individual claims).
- Algorithms were used that automatically accounted for interaction terms.

And the top 10 features for each group were different (Supplemental Material 4):

- Age: Age was relevant for all groups but one. For Blacks, it failed to make the Top 10.
- Race: Race & Ethnicity was the second most important factor to predict an adverse health event for Blacks. For other races and ethnicities, it did not make the top 10.
- Dual Eligible: This was a top 10 factor for all groups except Whites.
- Days Since Most Recent PCP Visit: This varied by Race & Ethnicity and was ranked differently between groups.

#### Reducing algorithmic unfairness

The Adverse Event Model was analyzed for fairness using Group Benefit Equality (GBE). ClosedLoop’s submission also demonstrated how GBE could be used to calculate and apply a threshold adjustment to potentially reduce disparities in program enrollment. The calculations in Table 2 are merely illustrative and not reflective of a particular program.

**Table 2.** Applying GBE as a threshold adjustment. Initial results showed most groups with GBE values above or close to 1.0. They also revealed two outlier groups, & #x201C;Asian” and & #x201C;Other & Unknown” whose GBE values indicated they would be underrepresented in a high-risk group.

| Population | Initial |  |  | Adjusted |  |  |  |
| --- | --- | --- | --- | --- | --- | --- | --- |
|  | Thres. | GBE | Population | Thres. | GBE | Population | Impact |
| Race / Ethnicity |  |  |  |  |  |  |  |
| Asian | 95% | 0.78 | 20,910 | 92.3% | 1.00 | 27,540 | +6,630 |
| Black | 95% | 1.25 | 327,114 | 95% | 1.25 | 327,114 | 0 |
| Hispanic | 95% | 1.07 | 57,069 | 95% | 1.07 | 57,069 | 0 |
| N Am Native | 95% | 1.31 | 20,298 | 95% | 1.31 | 20,298 | 0 |
| Oth / Unknown | 95% | 0.70 | 26,112 | 93.7% | 1.00 | 38,352 | +12,240 |
| White | 95% | 0.95 | 1,421,880 | 95% | 0.95 | 1,421,880 | 0 |
| Female Gender |  | 1.02 | 1,082,526 |  | 1.02 | 1,091,502 | +8,976 |
| Disabled<br>Individuals |  | 1.28 | 911,829 |  | 1.28 | 919,938 | +8,109 |
| Dual Eligible |  | 1.35 | 842,418 |  | 1.35 | 853,383 | +10,965 |
| Population Density |  |  |  |  |  |  |  |
| Urban Center |  | 0.99 | 271,728 |  | 1.00 | 273,819 | +2,091 |
| Rural |  | 0.97 | 13,770 |  | 0.97 | 13,821 | +51 |
| Full Population |  |  | 1,873,383 |  |  | 1,892,253 | +18,870 |

Calculations also illustrated using GBE to adjust the enrollment threshold on a per group basis. For example, attaining a GBE of 1.0 for the Asian group required lowering the enrollment threshold from 95% to 92.3%, which resulted in an additional 6,630 Asians being prioritized for care management. To ensure such a correction would not have significant unintended negative consequences, GBE measurements were computed for the adjusted model. After correction, no groups were significantly below 1.0.

## Discussion

ClosedLoop used the framework published by Paulson & Kent to evaluate their models for algorithmic bias and demonstrate how fairness criteria could be used to mitigate unfairness. To the authors’ knowledge, this is the first-ever application of a comprehensive framework to measure algorithmic bias and fairness in predictive models built using Medicare claims data, and the first demonstration of how fairness measures can facilitate adjustments to care management strategies to promote fairness. Also, to the authors’ knowledge, it is the only healthcare-specific data science platform to incorporate this capability.

The results of this application highlighted the importance of the framework’s distinct steps.

1. Representative samples: The data reflected Medicare members covered between 2013-2017, but not necessarily other populations. Thus, models developed for the Challenge should not be applied to other populations (e.g., Medicare Advantage, Medicaid, Commercial, etc.) or to other traditional Medicare populations outside the period 2013-2017 without further validation.
2. Subgroup validity: Calibration was less than 0.001 for all groups and AUC is within 3.2% of the population average for all groups. North American Natives had somewhat lower performance, which was likely because this subgroup had very few beneficiaries in the CMS data. Training on a larger cohort would be expected to improve performance for this group.
3. Label choice bias: This can arise whenever the desired outcome for prediction is different from the label on which an algorithm is trained, which is often a proxy. In 2019, Obermeyer et al. identified racial bias in an algorithm widely used to identify high-risk members based on medical costs.[5] Blacks tended to have lower costs for the same conditions as Whites. Using cost as a proxy for health status led to Blacks being identified as lower risk than Whites with similar conditions, an example of & #x201C;label choice” bias. This analysis showed the composite label was appropriate for population health interventions. However, it was also observed that the inclusion of Emergency Department (ED) visits might create label choice bias and should be considered based on the intervention. In real world settings, assessing label choice bias requires domain experience and cannot be automated. For models being developed for use in practice, it is important to align predictions with specific intervention programs, since better alignment of predictions with its intended use can reduce label choice bias.[6]
4. Feature bias and diversity: The models were developed from 1000s of candidate features. The final models included 753 features derived from claims and publicly available datasets. Using relevant and diverse features helped achieve high accuracy and validity across different subgroups.[12] Healthcare costs were not used in the model. Race / Ethnicity and Medicaid status were included, and the analysis showed their inclusion would likely reduce health disparities. Feature bias can arise due to differing healthcare behaviors or utilization thresholds, differences in access to care, differences in how diagnoses are ascertained, and data & #x2018;missingness’.[13] When evaluating whether to include race in the development or application of clinical algorithms, ClosedLoop addressed three questions recommended by experts [11]:
  a. **Was the need to include race based on robust evidence and statistical analyses?** Yes. Models that excluded race, Medicaid status, and SDoH factors introduced bias via subgroup invalidity and disproportionately lowered accuracy for Blacks and Hispanics relative to Whites and Other. Removing those variables was detrimental to all races; accuracy decreased for all models when race was excluded. (Supplemental Material 5).
  b. **Was there a plausible causal mechanism for the inclusion of race?** Yes. It has been broadly established that health is strongly influenced by a matrix of socio-ecological influences, including race and racism, which shape and perpetuate health inequities.[3-5]
  c. **Would including race in a model relieve or exacerbate health inequities?** Removing race and other Social Determinants of Health (SDoH) variables introduced bias that could exacerbate health inequities. Specifically, model results showed that Whites and Asians would be enrolled in care management at the expense of Blacks, Hispanics, and North American Natives. Dual-eligible beneficiaries might also receive fewer resources (Supplemental Material 5).
5. Algorithmic unfairness: Reducing algorithmic bias may be insufficient to address fairness concerns in polar decision contexts. In such cases, it is possible for a model to be algorithmically unbiased yet still result in decisions deemed unfair.[13] ClosedLoop’s submission included an example of implementing a fairness adjustment by changing thresholds for certain groups. For models being designed for use in real world situations, the decision to apply a fairness adjustment should be made by all stakeholders and consider the specific decisions being made.

A strength of the submission was its ability to demonstrate precision and transparency in the comprehensive measurement of algorithmic bias from multiple sources, including representativeness of data, subgroup validity, label choice, and feature bias. For feature bias, the submission made a detailed examination of feature selection and diversity, including evaluating the appropriateness of including race in the development of the algorithms.

Another strength was the submission’s demonstration of the importance of using fairness metrics appropriate for healthcare. Differing notions of fairness are appropriate based on context which has led to the development of different fairness metrics.[14] Some are inappropriate for healthcare settings where observed rate differences can either be expected and appropriate (e.g., mammograms by gender) [15] or the very issue under scrutiny (e.g., pregnancy-related death by race). For example, Disparate Impact is one of the most commonly-applied metrics outside healthcare. It requires each class to have the same predicted event rate. A naive application in healthcare leads to nonsensical results that could increase health inequities (e.g., forcing both men and women to have equal risk of breast cancer). Another example is Disparate Treatment, a causal notion of fairness that requires similar individuals not be treated differently solely because they are in a different group, but generally cannot be assessed in observational data. And Equality of Opportunity metrics, while they attempt to balance a model’s predictive power across groups and account for observed differences in event rates, fail to quantify unfairness and are not comparable across settings.

A weakness of the submission was that it did not assess other important groups for algorithmic bias and fairness. Research has shown that health disparities and vulnerability to bias and fairness also affects groups that are distinct for other reasons (e.g., data collection challenges, unique health considerations) [16-18]. Some of these groups were excluded from the datasets (e.g., veterans) while others were not explicitly identified within the data (e.g., LGBT individuals). In addition, the examination of race and ethnicity was confined to the definitions within the database.

Also, ClosedLoop did not evaluate the 12-Month Mortality Model for label choice bias. It was assumed that model would inform a non-polar decision. It is notable, however, that mortality can be subject to label choice bias if used for decision making and Advanced Care Planning when observed mortality is materially different than mortality under ideal care.[13]

## Conclusion

Algorithms play an increasingly central role in healthcare. Their use in value-based care must address their potential to perpetuate health disparities. It is possible for healthcare organizations to adopt policies and practices that enable them to design, implement, and maintain algorithms that are highly accurate, unbiased, and fair.

## Methods

ClosedLoop applied a comprehensive framework for assessing algorithmic bias and fairness. They also developed and applied a metric appropriate for real-world healthcare settings capable of being used to assess and reduce the presence and impact of unfairness.

### A comprehensive framework for assessing algorithmic bias and fairness

A comprehensive framework by Paulus & Kent was used to examine algorithmic bias and fairness (Fig. 2).[13] Their framework emphasizes concepts crucial to assessing bias in healthcare, particularly that fairness concerns are not equivalent across prediction-supported decisional contexts, and bias and fairness are distinct and must be addressed separately.

**Fig 2.**
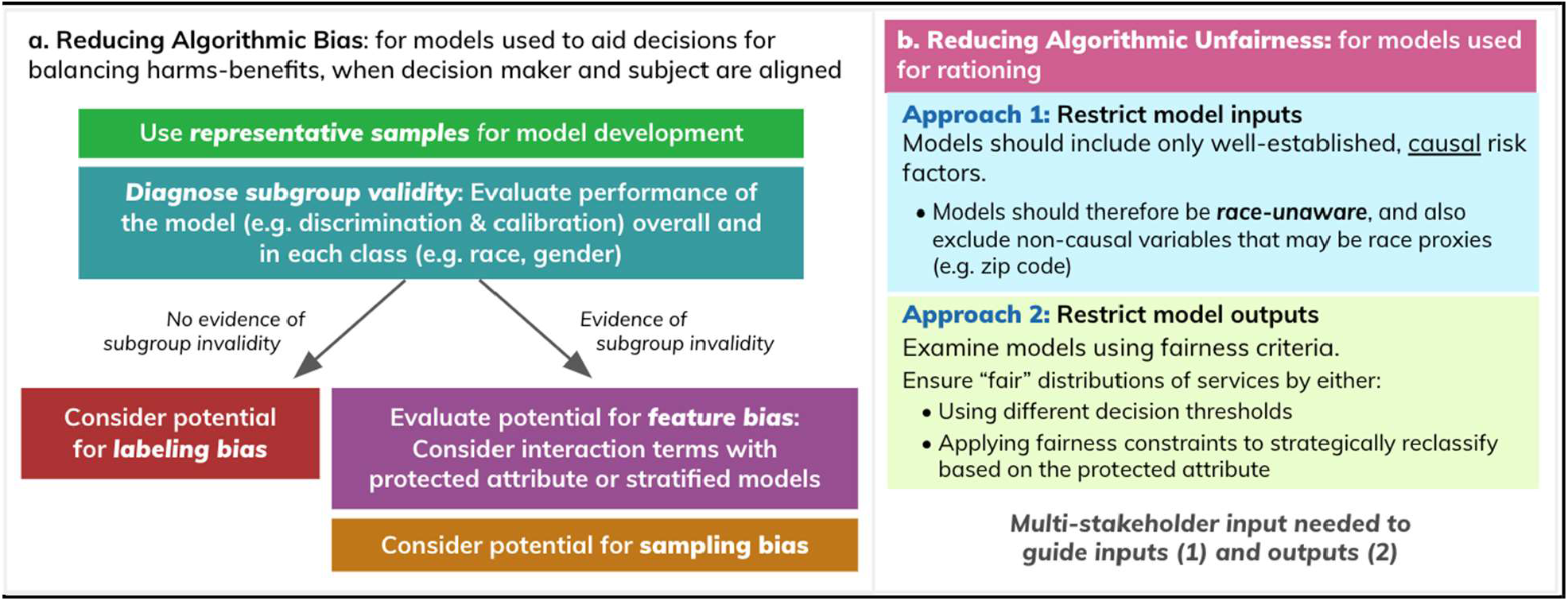
Framework for examining bias and fairness.

- Polar decisions: When the goals of everyone in the decisional context are not necessarily aligned and potentially conflicting claims could be advanced (e.g., when predictions are used to prioritize patients to limited resources which can improve health)
- Non-Polar decisions: When interests are aligned and parties to the decision share a common purpose (e.g. the goal of accurate prognoses, as in shared decision-making when patients and physicians balance the benefits and harms of specific care choices).
- Bias: Related to model design, data, and sampling that may disproportionately affect model performance in a certain subgroup.
- Fairness: Related to applying model results. Reducing bias and differential performance may be insufficient to address fairness concerns in polar decisions.

In Stage 1, participants were allowed to submit different models. ClosedLoop’s submission was a 30-day Adverse Events Model. This was a composite model with predictions for 12 speci?c events and a composite prediction reflecting the risk of any of those events occurring within 30 days (Supplemental Material 6). In Stage 2, all participants submitted a model to predict 12-month all-cause mortality.

In applying the framework, ClosedLoop assumed the 30-Day Adverse Event Model would be used to identify patients for care management (a polar decision), while the 12-Month Mortality Model would be used to catalyze patient-physician discussions regarding serious illness care goals and Advance Care Planning (a non-polar decision).

### A New Fairness Metric for Healthcare: Group Benefit Equality

ClosedLoop developed and applied a metric for assessing fairness in healthcare settings called Group Benefit Equality (GBE).[19] GBE assesses the ratio of predicted risk to actual risk, where predicted risk is the total of true positives (TP) and false positives (FP), and actual risk in the total of true positives (TP) and false negatives (FN). In this way, GBE assess whether groups are being allocated resources in proportion to their observed need.[1]

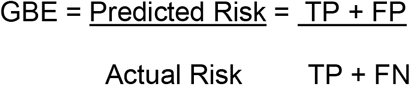

Like another fairness metric, Equality of Opportunity, GBE defines criteria a model must meet in order to be considered fair. But Equality of Opportunity provides no guidance on the degree to which a model is unfair when it fails to satisfy the criteria; GBE does.

Specifically, GBE provides a standardized measure of the degree to which a model is fair or unfair that can be applied consistently in different situations. GBE calculates a value for each protected group that equates to 1.0 when that group is treated fairly (i.e., when its predicted risk equals its actual risk), which varies (up or down) to reflect the degree of over/under-allocation of resources that would occur and quantifies any necessary threshold adjustments for program enrollment.

This article uses the SQUIRE reporting guidelines.[20]

## Data Availability

The datasets provided by CMS for the AI Health Outcomes Challenge were only supplied to participating contestants. ClosedLoop is not legally able to publicize the datasets. Comparable datasets to the ones used in the challenge are publicly available through CMS.

## Acknowledgements

ClosedLoop would like to thank the following collaborators for their contributions:

- Dr. David Kent from Tufts Predictive Analytics and Comparative Effectiveness (PACE) Center for providing guidance on how to apply his bias and fairness framework to the problems in this challenge.[13]
- Dr. Ziad Obermeyer, Emily Bembeneck and Michael Stern from UC Berkeley & the Chicago Booth Center for Applied Artificial Intelligence for reviewing our approach to label choice bias.[6]
- Booz Allen Hamilton - technical and fairness policy consulting and review.
- The Office of Senator Cory Booker for discussing our proposed approach to addressing bias and fairness.

In addition, authors would like to acknowledge Shaayaan Sayed and Brett Castellanos for their contributions to conception and design, evaluation of data, creation of models, and application of a comprehensive framework for assessing bias and fairness.

## Supporting information

**S1 Fig. Assessment of label choice bias**. Fig 1

**S2 Fig. Framework for examining bias and fairness**. Fig 2

**S1 Table. Data representativeness**. Table 1

**S2 Table. Applying GBE as a threshold adjustment**. Table 2

## Ethical statement

The authors are accountable for all aspects of the work in ensuring that questions related to the accuracy or integrity of any part of the work are appropriately investigated and resolved. All procedures performed in this submission were in accordance with the Declaration of Helsinki (as revised in 2013). Because this research was conducted using de-identified, anonymized, and obfuscated data and was conducted in compliance with the ethical requirements of the Centers for Medicare and Medicaid Services as part of the AI Health Outcomes Challenge no informed consent or ethical review was required.

## References

1. Pollack CE. Accountable Care Organizations and Health Care Disparities. JAMA. 2011 apr 27;305(16):1706.

2. Werner RM. Does Pay-for-Performance Steal From the Poor and Give to the Rich? Annals of Internal Medicine. 2010 sep 7;153(5):340.

3. Syed M, Mehmud. Nontraditional Variables in Healthcare Risk Adjustment Society of Actuaries’ Health Section [Internet]. 2013. Available from: https://www.soa.org/globalassets/assets/files/research/projects/research-2013-nontrad-var-health-risk.pdf

4. Wennberg DE, Sharp SM, Bevan G, Skinner JS, Gottlieb DJ, Wennberg JE. A population health approach to reducing observational intensity bias in health risk adjustment: cross sectional analysis of insurance claims. BMJ. 2014 apr 10;348(apr10 2):g2392–2.

5. Obermeyer Z, Powers B, Vogeli C, Mullainathan S. Dissecting racial bias in an algorithm used to manage the health of populations. Science [Internet]. 2019 oct 25;366(6464):447–53. Available from: https://science.sciencemag.org/content/366/6464/447.full

6. Obermeyer Z, Nissan R, Stern M, Eaneff S, Bembeneck E, Mullainathan S. Algorithmic Bias Playbook [Internet]. 2021 [cited 2021 Jul 21]. Available from: https://www.chicagobooth.edu/-/media/project/chicago-booth/centers/caai/docs/algorithmic-bias-playbook-june-2021.pdf

7. Booker (D-NJ) and Wyman (D-OR). Letter to CMS Regarding Disturbing Revelations of Flawed Algorithms Impacting Care for Black Patients [Internet]. Scribd. 2019 [cited 2021 Jul 23]. Available from: https://www.scribd.com/document/437955444/Booker-Wyden-CMS-Letter

8. Centers for Medicare and Medicaid Services. Artificial Intelligence (AI) Health Outcomes Challenge | CMS Innovation Center [Internet]. innovation.cms.gov. 2021 [cited 2021 Jul 23]. Available from: https://innovation.cms.gov/innovation-models/artificial-intelligence-health-outcomes-challenge

9. ClosedLoop.ai. How ClosedLoop Won the CMS AI Challenge [Internet]. www.closedloop.ai. 2021 x[cited 2021 Oct 4]. Available from: https://www.closedloop.ai/resources/how-closedloop-won-the-cms-ai-challenge

10. Matthews BW. Comparison of the predicted and observed secondary structure of T4 phage lysozyme. Biochimica et Biophysica Acta (BBA) - Protein Structure. 1975 oct;405(2):442–51.

11. Vyas DA, Eisenstein LG, Jones DS. Hidden in Plain Sight — Reconsidering the Use of Race Correction in Clinical Algorithms. Malina D, editor. New England Journal of Medicine. 2020 jun 17;383(9).

12. Kasthurirathne SN, Grannis S, Halverson PK, Morea J, Menachemi N, Vest JR. Precision Health–Enabled Machine Learning to Identify Need for Wraparound Social Services Using Patient- and Population-Level Data Sets: Algorithm Development and Validation. JMIR Medical Informatics. 2020 jul 9;8(7):e16129.

13. Paulus JK, Kent DM. Predictably unequal: understanding and addressing concerns that algorithmic clinical prediction may increase health disparities. npj Digital Medicine. 2020 jul 30;3(1).

14. Verma S, Rubin J. Fairness definitions explained. Proceedings of the International Workshop on Software Fairness - FairWare ’18 [Internet]. 2018; Available from: https://fairware.cs.umass.edu/papers/Verma.pdf

15. Anderson WF, Jatoi I, Tse J, Rosenberg PS. Male Breast Cancer: A Population-Based Comparison With Female Breast Cancer. Journal of Clinical Oncology. 2010 jan 10;28(2):232–9.

16. Agency for Healthcare Research and Quality. 2019 National Healthcare Quality and Disparities Report [Internet]. www.ahrq.gov. Rockville, MD: AHRQ; 2019. Available from: https://www.ahrq.gov/research/findings/nhqrdr/nhqdr19/index.html

17. National Academies of Sciences, Engineering and Medicine. Communities in Action [Internet]. Weinstein JN, Geller A, Negussie Y, Baciu A, editors. Washington, D.C.: National Academies Press; 2017. Available from: https://www.nap.edu/read/24624/chapter/7

18. Berwick DM. Choices for the “New Normal.” JAMA [Internet]. 2020 Jun 2 [cited 2022 Sep 19];323(21):2125. Available from: https://jamanetwork.com/journals/jama/fullarticle/2765699

19. Gartner J. A New Metric for Quantifying Machine Learning Fairness in Healthcare [Internet]. www.closedloop.ai. 2020 x[cited 2021 Jul 21]. Available from: https://www.closedloop.ai/post/a-new-metric-for-quantifying-machine-learning-fairness-in-healthcare

20. Ogrinc G, Davies L, Goodman D, Batalden P, Davidoff F, Stevens D. SQUIRE 2.0 (Standards for QUality Improvement Reporting Excellence): revised publication guidelines from a detailed consensus process: Table 1. BMJ Quality & Safety. 2015 sep 14;25(12):986–92.

## Table 1 References

[dataset] a. Data from: American Community Survey: 2015–2019 ACS 5-Year Data Profile. United States Census Bureau. 2020. https://www.census.gov/acs/www/data/data-tables-and-tools/data-profiles/ (accessed 21 Jul 2021).

[dataset] b. Data from: Americans With Disabilities: 2010 United States Census Bureau. (updated 2017). https://www.census.gov/data/tables/2010/demo/disability/p70-131.html (accessed 21 Jul 2021).

c. Data Analysis Brief: Medicare-Medicaid Dual Enrollment 2006 through 2018. Centers for Medicare and Medicaid Services. 2019. https://www.cms.gov/Medicare-Medicaid-Coordination/Medicare-and-Medicaid-Coordination/Medicare-Medicaid-Coordination-Office/DataStatisticalResources/Downloads/MedicareMedicaidDualEnrollmentEverEnrolledTrendsDataBrief2006-2018.pdf (accessed 21 Jul 2021).

[dataset] d. Data from: 2013 Rural-Urban Continuum Codes. United States Department of Agriculture. (updated 2020). https://www.ers.usda.gov/data-products/rural-urban-continuumcodes.aspxcodes.aspx (accessed 21 Jul 2021).

